# Baseline analysis of patients presenting for surgical review of ACL rupture reveals heterogeneity in patient-reported outcome measures

**DOI:** 10.1101/2020.03.08.20020990

**Authors:** Chee Han Ting, Corey Scholes, David Zbrojkiewicz, Christopher Bell

## Abstract

Despite establishment of successful surgical techniques and rehabilitation protocols for ACL reconstruction, published return to sport rates are less than satisfactory. This has led orthopaedic surgeons and researchers to develop more robust patient selection methods and investigate prognostic patient characteristics. No previous studies have integrated baseline characteristics and responses to PROMs of patients with ACL rupture presenting for surgical review.

Patients electing to undergo ACL reconstruction under the care of a single orthopaedic surgeon at a metropolitan public hospital were enrolled in a clinical quality registry. Patients completed VR-12 PCS and MCS scores, Tegner activity scale and IKDC questionnaires at presentation. Total scores were extracted from the electronic registry, and a machine learning approach (k-means) was used to identify subgroups based on similarity of questionnaire responses. The average scores in each cluster were compared using ANOVA (Kruskal-Wallis) and nominal logistic regression was performed to determine relationships between cluster membership and patient age, gender, BMI and injury-to-examination delay.

A sample of 107 patients with primary ACL rupture were extracted, with 97 (91%) available for analysis with complete datasets. Four clusters were identified with distinct patterns of PROMs responses. These ranged from lowest (Cluster 1) to highest scores for VR-12 and IKDC (Cluster 4). In particular, Cluster 4 returned median scores within 6 points of the PASS for the IKDC score for ACL reconstruction (70.1, IQR 59 - 78). Significant (p<0.05) differences in PROMs between clusters was observed using ANOVA, with variance explained ranging from 40-69%. However, cluster membership was not significantly associated with patient age, gender, BMI or injury-to-examination delay.

Patients electing to undergo ACL reconstruction do not conform to a homogenous group but represent a spectrum of knee function, general physical and mental health, and preinjury activity levels, which may not lend itself to uniform treatment and rehabilitation protocols. The factors driving these distinct responses to PROMs remain unknown, but are unrelated to common demographic variables.

## Introduction

A decision to undergo anterior cruciate ligament (ACL) reconstruction requires careful consideration of a patient’s physical characteristics, functional demands and lifestyle ^1^. The aim of surgery is to restore stability to the patient’s knee to facilitate participation in a postoperative rehabilitation program that prepares them for return to preinjury activities and sport. Despite the establishment of successful surgical techniques and rehabilitation protocols, published return to sport rates are less than satisfactory ^2^. Further evidence from randomised controlled trials has not clearly identified those patients who will benefit from ACL reconstruction and those who will not ^3^. This has led orthopaedic surgeons and researchers to focus on developing more robust patient selection methods and to investigate patient characteristics that predict treatment outcomes ^4^.

Outcome assessment in ACL reconstruction continues to evolve from traditional clinician-based physical measurements towards patient-reported outcome measures (PROMs) and participation-based outcomes such as return to sport ^5,6^. PROMs are clinically validated tools that are used to measure the impact of knee injury and treatment on patients’ function and overall health ^5^. They can be implemented in different ways to assess the effect of interventions and baseline variables on the efficacy of treatment. When collected pre- and postoperatively, PROMs can be used to detect clinically significant changes that reflect what is a meaningful outcome for the patient ^7^. Recent studies have also investigated utilising preoperative PROMs to predict postoperative outcomes in orthopaedic surgery, highlighting the importance of understanding the baseline characteristics of patients ^4,8^.

There is a growing body of literature reporting the use of preoperative patient factors and PROMs assessed by multivariable analysis to predict outcomes in ACL reconstruction ^4,9,10^. Previous work has identified several candidate predictors, but no studies have integrated the baseline characteristics and responses to PROMs of patients with ACL rupture presenting for surgical review ^10^. Compared to multivariable regression analysis, a machine learning technique such as cluster analysis provides an opportunity to assign patients to subgroups based on similar baseline characteristics and responses to multiple PROMs. This method of analysis incorporates preoperative patient assessment across multiple domains and acknowledges the importance of psychological factors on the outcome of ACL reconstruction ^11–13^.

Cluster analysis of patients presenting for surgical assessment of ACL rupture may help identify those who are expected to do poorly with surgery. Such patients could be targeted by specific interventions aimed at improving their outcomes ^4,13^. In addition, not all patients with ACL rupture need to have surgery and knowledge of subgroups may define a clearer role for nonoperative management ^3^. The aim of this study was to identify subgroups of patients enrolled in a clinical quality registry based on their responses to multiple PROMs at the time of diagnosis and decision to undergo ACL reconstruction. We hypothesised that patients presenting for surgical review of ACL rupture would belong to distinct subgroups that represent a range of functional deficits, general health, preinjury activity level and responses to PROMs.

## Methods

We conducted a cross-sectional analysis embedded within an institutional registry. Patients electing to undergo ACL reconstruction under the care of a single orthopaedic surgeon at a metropolitan public hospital were enrolled in a clinical quality registry. The Shoulder, Hip, Arthroplasty and Knee Surgery (SHARKS) clinical quality registry was implemented in June 2017 to pilot with one surgeon within the department for shoulder and knee procedures and was expanded in April 2019 to include three additional surgeons for hip and knee procedures, including ACL reconstruction (ACTRN 12617001161314). Patients presenting with ACL rupture were seen by house officers and registrars under the supervision of one of the participating consultant surgeons in the outpatient department and extracted for further analysis. Diagnosis and decision to undergo ACL reconstruction was made during the consultation. Prior to completion of the consultation, patients were asked to participate in the registry on an opt-in basis and their enrolment was confirmed by written informed consent. Ethical approval for the registry was granted by the local health district HREC (HREC/16/QPAH/732).

Exclusion criteria in the present analysis were i) diagnosis of ACL rupture associated with knee dislocation or requiring multiligament reconstruction, ii) revision ACL reconstruction, iii) missing PROMs responses. Preoperative Veterans RAND 12-item Health Survey (VR-12) Physical Component Summary (PCS) and Mental Component Summary (MCS) scores, Tegner activity scale and International Knee Documentation Committee (IKDC) Subjective total scores were collected along with baseline demographic data on paper forms following outpatient consultation. These forms were then electronically scanned for data entry and storage in dedicated database software (Socrates v3.5, Ortholink Pty Ltd, Aus).

### Statistical analysis

A dataset of all eligible patients presenting between June 2017 and April 2019 was extracted from the registry database following routine quality auditing for completeness and validity. Selection and reporting bias were mitigated by using census sampling within the registry and a high proportion of complete datasets within the sample. Age at initial examination (years) was calculated from the date of birth and date of examination. The interval between the date of injury and date of examination was also calculated (weeks). Baseline characteristics of the included sample were summarised using median and interquartile ranges. Categories were calculated for BMI using Australian Bureau of Statistics definitions ^14^ and the patient acceptable symptom state (PASS) for the IKDC for ACL reconstruction threshold of 75.9 ^15^. PROMs scores were assessed for normality with Anderson-Darling normality tests and visualised with probability plots with 95% confidence intervals. An unsupervised learning approach (k-means clustering) was used to identify subgroups based on the similarity of VR-12, Tegner activity scale (pre-injury) and IKDC responses. An initial cluster number (k) of 4 was set based on a pilot analysis and the PROMs scores standardised (sample mean subtracted from the individual score and divided by the sample standard deviation). A post-hoc validation of the initial k selected was performed by repeating the analysis for different *k* between 2 and 10 and examining silhouette plots for misclassification rates (supplementary material). Kruskal-Wallis ANOVAs with multiple comparisons based on Dunn’s test were used to assess differences between the average PROMs score in each cluster. Eta-squared (η^2^) was calculated as the ratio between the sum of squares of the cluster effect and the total sum of squares to describe the effect size for each ANOVA. Ordinal logistic regression was performed to determine the relationship between cluster membership and patient age, gender, BMI and injury to examination time. A post-hoc chi-squared analysis was performed on the distribution of responses to Question 7 of the IKDC relative to cluster membership. All statistical analyses were performed in Minitab (v18, Minitab Inc, MA, USA), with alpha set at 5% for all tests.

## Results

### Baseline characteristics

A sample of 107 patients with primary ACL rupture were extracted, with 97 (91%) available for analysis with complete datasets (Figure 1). Baseline characteristics of the sample are summarised in Table 1. The sample included a high proportion of males (65%) and of overweight-obese patients (59%) classified by BMI, with a proportion (8.2%) returning IKDC scores preoperatively that exceeded the PASS threshold. The VR-12 and Tegner activity scale responses did not follow normal distributions, whereas the IKDC did (Figure 2).

**Table 1:** Baseline characteristics of patients included for analysis

|  | **N** | **Median (IQR)** | **Classifications (%)** |
| --- | --- | --- | --- |
| **Age** | 97 | 24.5 (21.2 - 29.9) |  |
| **Date of injury to examination (weeks)** | 87 | 5.9 (2.9 - 10.8) |  |
| **BMI** | 102 | 26.7 (24.6-30.3) | Normal: 30.4<br>Overweight: 41.2<br>Obese: 28.4 |
| **VR-12 PCS** | 97 | 41.5 (32.9-47.9) |  |
| **VR-12 MCS** | 97 | 53.4 (41.3-59.6) |  |
| **Tegner activity scale preinjury** | 97 | 9 (6.5 - 9) |  |
| **Tegner activity scale preoperative** | 91 | 3 (2 - 4) |  |
| **IKDC** | 95 | 49.4 (37.4 - 62.1) | >PASS: 8.4 |

**Figure 1:**
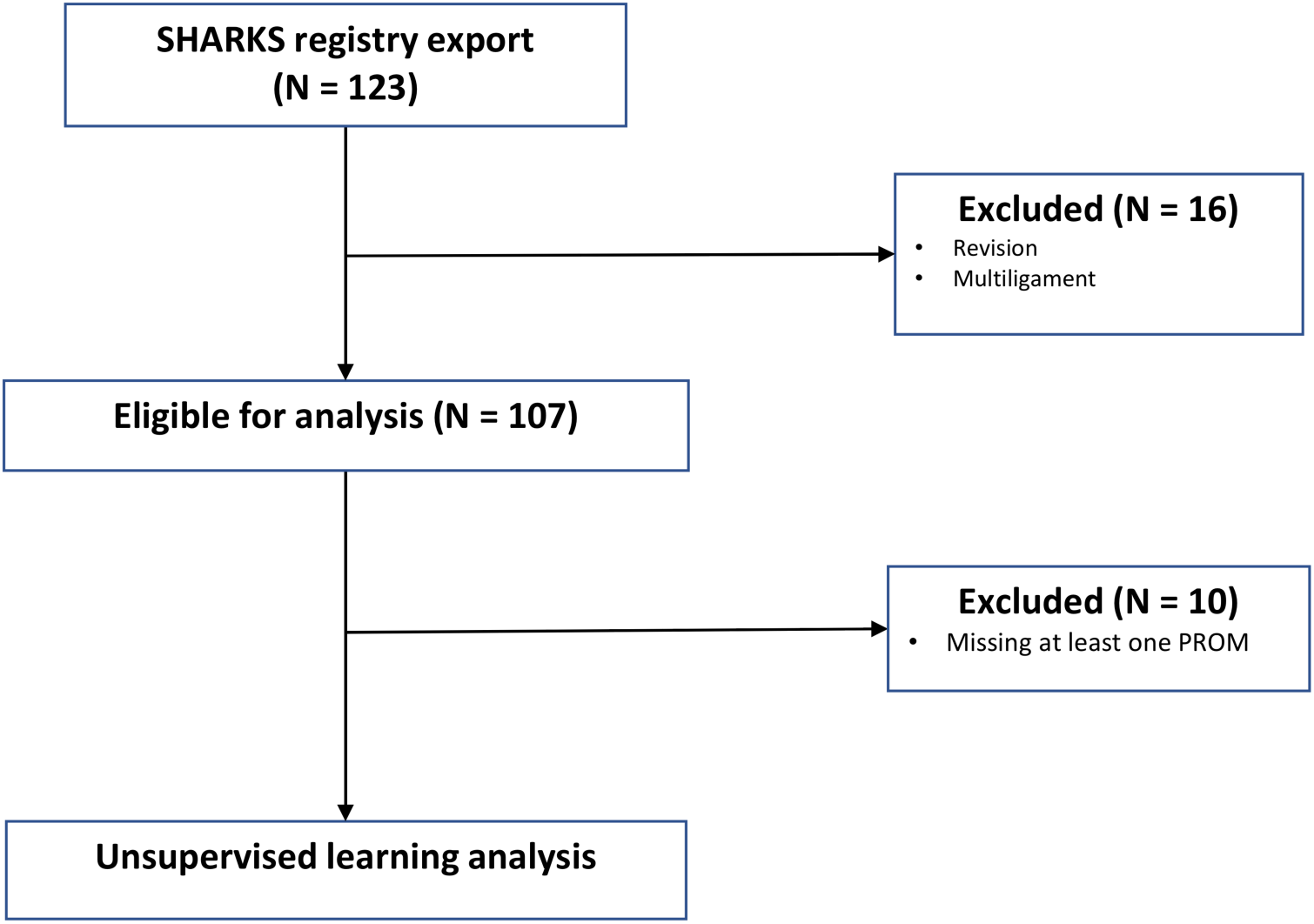
STROBE ^16^ flow diagram illustrating extraction from the SHARKS registry and analysis

**Figure 2:**
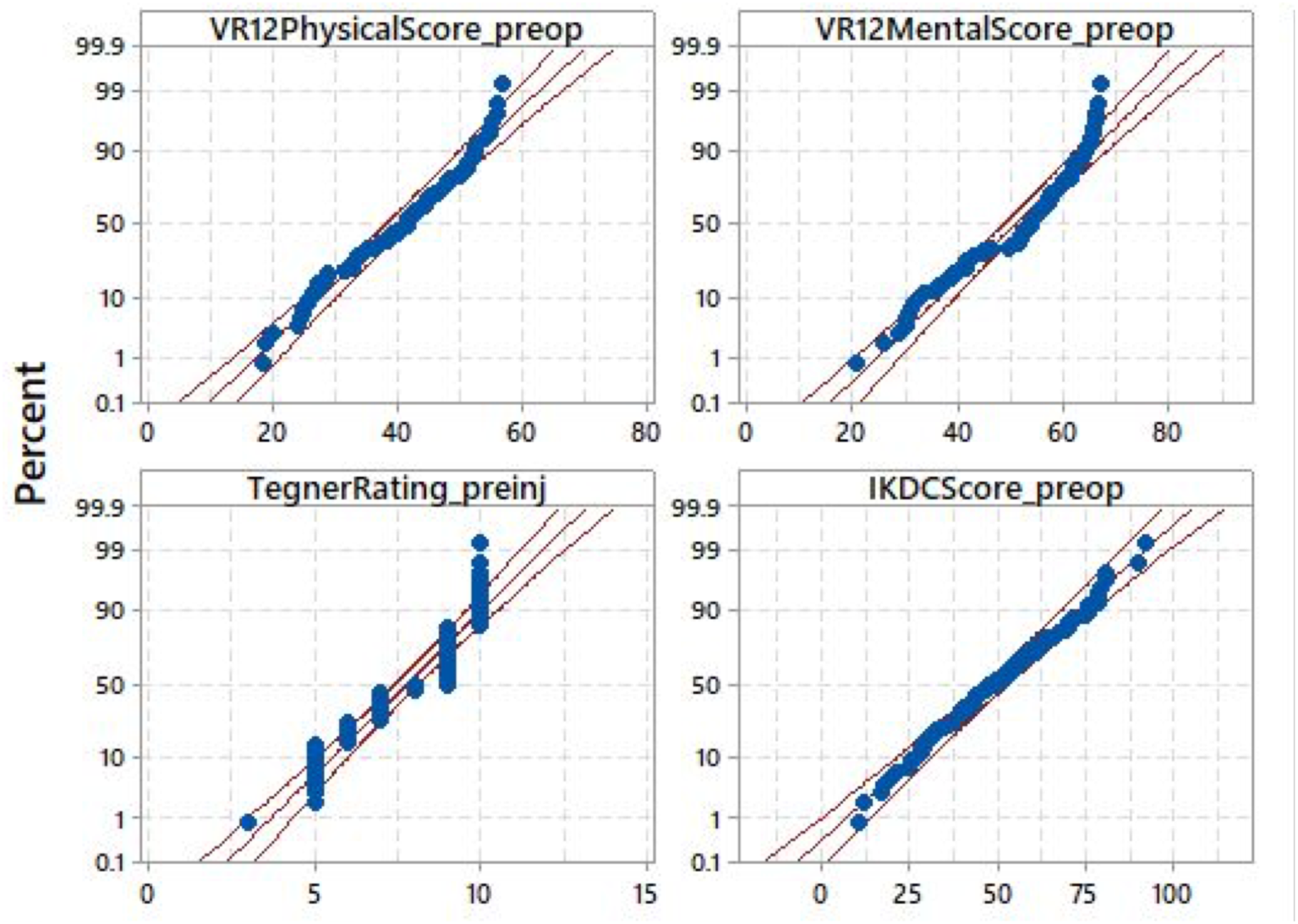
Distributions of preoperative patient-reported outcomes measure (PROMs) scores.

### Cluster analysis

Four subgroups (Clusters 1 to 4) were identified with distinct patterns of responses to the PROMs (Figure 3). They ranged from the lowest scores (Cluster 1) to the highest scores for VR-12 and IKDC (Cluster 4). Significant (p<0.05) differences in PROMs between clusters was observed, with variance explained ranging from 40 to 69% (Table 2). However, cluster membership was not significantly associated with patient age, gender, BMI or injury to examination delay (Table 3). Post-hoc analysis of responses to Question 7 of the IKDC *“What is the highest level of activity you can perform without significant giving way in your knee?”* revealed a higher rate of patients in Cluster 1 (low scores) unable to perform any activities, compared to Cluster 4 (high scores) with lower rates of patients responding with light activities and a higher rate responding with moderate-strenuous (Table 4).

**Table 2:** Significant differences between clusters for component scores

| Cluster | 1 | 2 | 3 | P-Value | $\eta^2$ (%) |
| --- | --- | --- | --- | --- | --- |
| **VR-12 PCS** | Vs 2,3,4 | Vs 3,4 |  | <0.001 | 55 |
| **VR-12 MCS** | Vs 2,3,4 | Vs 4 | Vs 4 | <0.001 | 40 |
| **Tegner** | Vs 2,3,4 | Vs 3 | Vs 4 | <0.001 | 52 |
| **IKDC** | Vs 2,3,4 | Vs 4 | Vs 4 | <0.001 | 69 |

**Table 3:** Logistic regression results for selected predictors for cluster membership

|  | Odds Ratio | 95%CI | P-value |
| --- | --- | --- | --- |
| **Age** | 1.09 | 0.99 - 1.2 | 0.069 |
| **Gender** | 0.37 | 0.09 - 1.54 | 0.174 |
| **BMI** | 1.07 | 0.91 - 1.26 | 0.395 |
| **Date of injury to examination** | 0.99 | 0.96 - 1.02 | 0.51 |

**Table 4:**
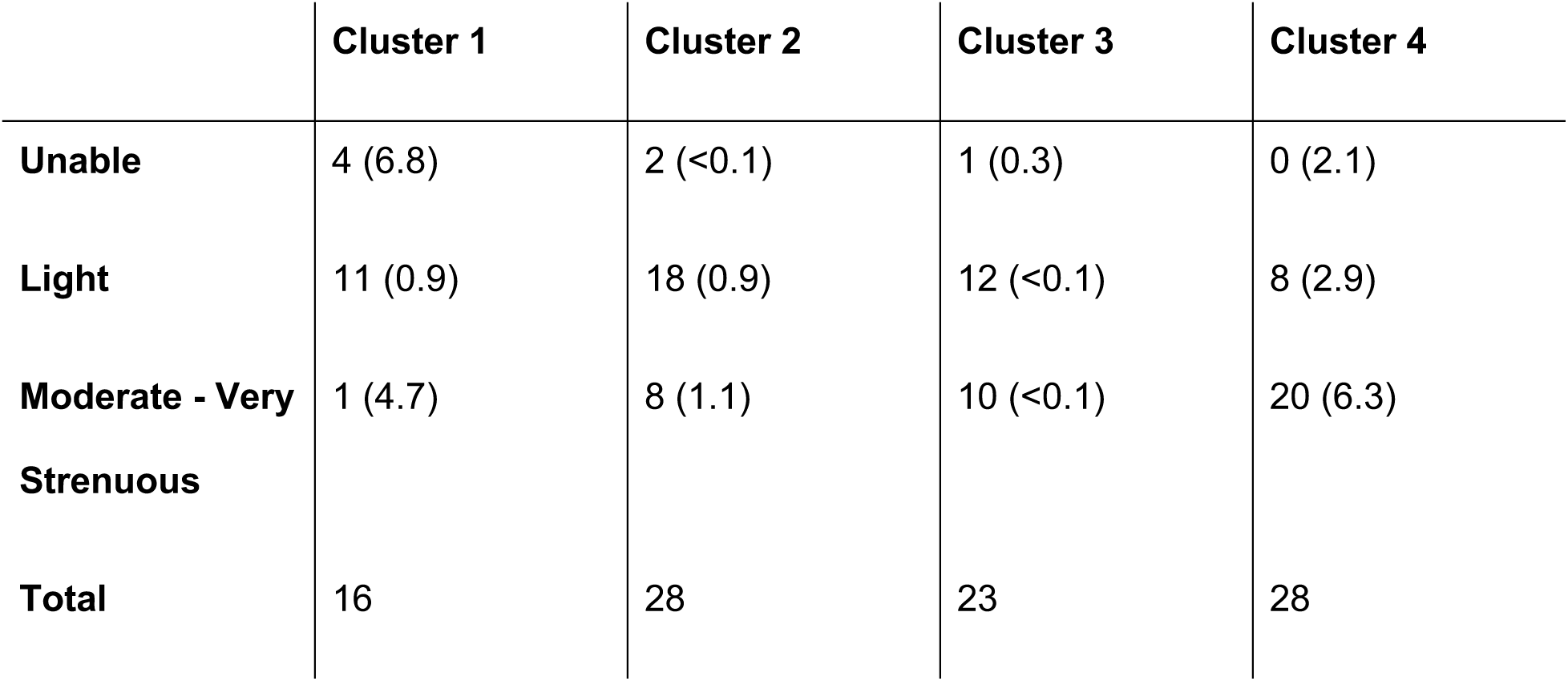
Number of responses in each IKDC Q7 (episodes of instability) category versus cluster membership (***X***^2^ contribution)

**Figure 3:**
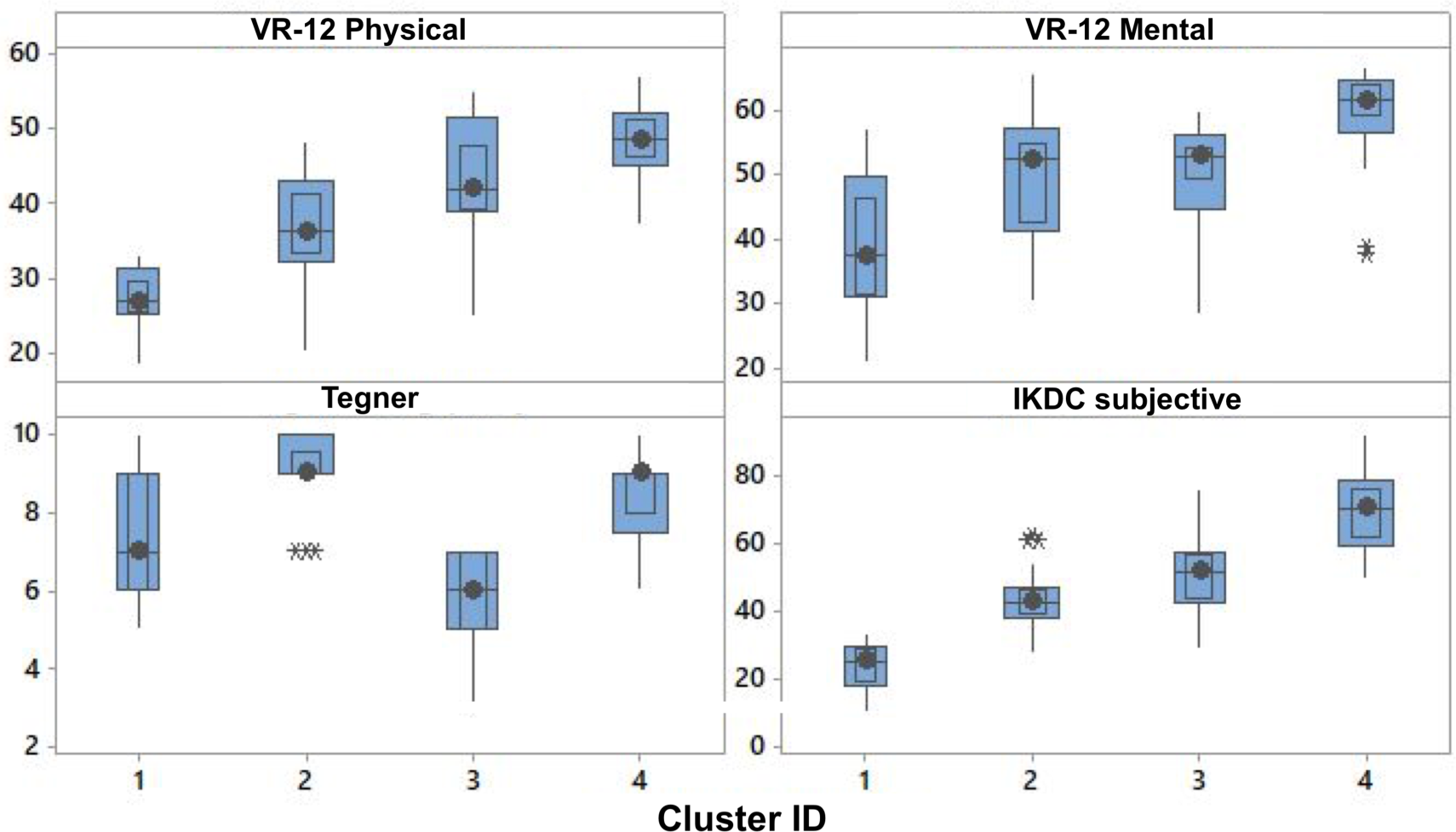
Median PROMs scores (solid circle), bounded by interquartile range boxes (blue box representing middle 50% of data) and whiskers (representing 75% of data) separated by cluster.* outliers

## Discussion

Our study is the first to analyse the baseline characteristics of patients undergoing ACL reconstruction across preinjury activity level, overall mental and physical health and knee function scores simultaneously. Patients presenting for ACL reconstruction belong to subgroups with distinct patterns of responses. We were able to identify four clusters, with one group having the lowest scores for VR-12 and IKDC and another having the highest scores for VR-12 and IKDC. Patients electing to undergo ACL reconstruction may present with knee function ranging from poor to normal. Cluster 4 returned average scores (median 70.1, IQR 59 - 78) that were within 6 points of the PASS for the IKDC score for ACL reconstruction ^15^. Our results also showed that membership to a particular cluster was not significantly associated with age, gender, BMI or time from injury to examination.

Post-hoc analysis of the IKDC scores revealed that Cluster 4, the subgroup with the highest overall health and knee function scores, reported the ability to perform a higher level of activity without experiencing instability, at a higher than expected rate compared to Cluster 1. These findings highlight the need to clearly establish the reasons why patients decide to undergo ACL reconstruction. Patients elect to undergo reconstruction without having experienced poor knee function ^17^. Decisions to have surgery are based on having a high preinjury activity level, the presence of concomitant meniscal or ligament pathology, an anticipation of future issues with the knee or the influence of high level athletes undergoing reconstruction ^17–19^. Feucht et al. reported that following ACL reconstruction, patients expect a normal functioning knee that allows a return to the same level of preinjury activity and without the risk of developing osteoarthritis in the future ^19^. Patients with high present self-efficacy and high future self-efficacy of knee function, which are indicators of high expectations for postoperative knee function, are associated with higher activity level, less knee symptoms and better physical performance after surgery ^20^. However, there may be multiple competing explanations for these findings. Firstly, the fulfilment of expectations rather than the experience of poor knee function and instability may be the reason why patients with high preoperative PROMs scores still opt to have surgery. Cluster 4 may represent this subgroup of patients with high expectations and positive psychological traits that lend themselves to better outcomes and return to sport rates. Secondly, the IKDC question may not be framed appropriately to resolve between current symptoms and previous activity level. Lastly, there may be as yet unknown factors influencing the relationship between cluster membership (baseline patient outcomes) and the decision to undergo reconstruction. Further work is required to elucidate these relationships in a larger sample.

Analysing the baseline psychological profile of patients with ACL rupture enables preoperative screening to identify patients at risk of a poor surgical outcome ^12,13,21,22^. The identification of clusters with distinct scores for VR-12 demonstrates the range of psychological states within this cohort. Cluster 1 may represent a subgroup at risk of unsatisfactory outcomes following surgery given they rated their overall health and knee function the poorest. A successful outcome from ACL reconstruction requires a sustained effort and adherence to intense rehabilitation, as well as overcoming the fear of reinjury in order to return to previous activity ^13^. Higher levels of self-efficacy, optimism and motivation are associated with better knee function scores and rates of return to sport ^12,13^. Patients identified as having a poor psychological profile for surgery and rehabilitation should be subject to specific interventions and treatment protocols aimed at optimising their outcomes. Psychological interventions that have been studied include cognitive behavioural therapy, guided imagery, counseling and positive self-talk, however, these have demonstrated inconsistent results ^22,23^. Future studies should focus on establishing interventions that address psychological readiness to achieve maximal outcomes ^12,13^. The best method of measuring these outcomes has not been well established ^4,7^.

Outcome reporting and prognostic modelling continues to mature in the field of orthopaedic surgery. There is no consensus regarding how best to measure outcomes or detect clinical improvements after ACL reconstruction. There is agreement that the success of ACL reconstruction is underpinned by a complex interplay of pathological, treatment, physical and psychological factors necessitating a multidimensional approach to evaluating and managing patients ^6^. Previous studies have used multivariable regression analyses to identify individual predictors of successful outcomes ^4,5,10^. Our cluster analysis represents an advancement toward identifying subgroups of patients within a disease entity using multiple interacting PROMs. The implications of this allow outlier individuals to be identified and targeted by tailored interventions to optimise their outcome.

There is next to no published guidance regarding the management and outcomes of ACL deficient patients with very high and very low preoperative knee function found in Cluster 4 and Cluster 1 of our study respectively. Establishing the baseline characteristics of patients undergoing ACL reconstruction is important for outcome prediction and developing prognostic tools to aid preoperative patient counselling ^5,9,10^. Patient stratification according to risk profile and prognostic modelling has been established in other medical specialties and could better inform treatment recommendations for ACL rupture ^24^. Fithian et al. classified patients as high, moderate or low risk based on their risk of having late ACL reconstruction, using preinjury activity level and knee laxity measurements as predictors ^18,25^. Patients were then allocated to an early operative treatment protocol or a nonoperative rehabilitation program with the option of converting to having a later reconstruction if they were not satisfied with their initial outcome ^25^. The algorithm demonstrated that low risk groups were less likely to require later reconstruction than their higher risk counterparts also receiving initial nonoperative treatment and that a delayed reconstruction was still an effective option ^25^. Preinjury activity level was not associated with cluster membership in our study and this challenges the notion that we can stratify patients according to this factor in isolation. Therefore, a more comprehensive stratification method, utilising preoperative factors across multiple domains is required. The method used in the present study could be incorporated into a treatment algorithm to help guide clinical decision making.

The interpretation of the present results is constrained by the following factors that impact on its generalisability. Firstly, the modest sample size of our study compromises the ability to draw definite conclusions from the results. However, previous systematic reviews on ACL rupture have reported on multiple studies publishing similar sample sizes ^2,26,27^. Secondly, the timing of PROMs collection after the decision to undergo ACL reconstruction in our study could influence individual scores. Despite this, timing PROMs collection after consultation has been shown to improve PROMs completion rates ^28,29^. Our study achieved a completion rate of 91%, well above the expected rate of 80% ^29^. Thirdly, the analytic technique used, cluster analysis, is characterised by a lack of prior knowledge of the underlying structure of the group being analysed ^30^. In addition, different clustering algorithms will generate different clustering solutions for any given dataset ^31^. Nevertheless, the results of this study are presented in the context of a validated model with 4-clusters, using a commonly employed technique (k-means) for exploratory analysis. Further work is required to replicate these findings in other populations. Lastly, the present study did not include patients undergoing nonoperative treatment for ACL rupture. Earlier studies have established a role for nonoperative treatment for specific subgroups of patients ^25,32^. Inclusion of nonoperatively managed patients would provide a more comprehensive description of the baseline characteristics of patients presenting with ACL rupture. Future studies should utilise larger sample sizes across multiple centres, include all patients with ACL rupture and examine cluster membership in relation to postoperative outcomes.

## Conclusion

Patients electing to undergo ACL reconstruction do not conform to a homogenous group but represent a spectrum of knee function, general physical and mental health and preinjury activity level, which may not lend itself to uniform treatment and rehabilitation protocols. The factors driving these distinct responses to PROMs remain unknown but are not related to common patient demographic variables. The identification of clusters, with patients presenting with very high and very low knee function may provide the basis for new models of care to allocate patients to individualised treatment protocols. However, further work is required to elucidate the decision to undergo surgery in this patient population, particularly in high-functioning patients.

## Data Availability

De-identified data will be available on reasonable request.

## Acknowledgements

The authors gratefully acknowledge Mac Cowley, Milad Ebrahimi, Andrew Klissanin and Ken Morrisey for their assistance in data collection, retrieval and analysis.

